# Percolation across households in mechanistic models of non-pharmaceutical interventions in SARS-CoV-2 disease dynamics

**DOI:** 10.1101/2021.06.07.21258403

**Authors:** Caroline Franco, Leonardo Souto Ferreira, Vítor Sudbrack, Marcelo Eduardo Borges, Silas Poloni, Paulo Inácio Prado, Lisa J White, Ricardo Águas, Roberto André Kraenkel, Renato Mendes Coutinho

## Abstract

Since the emergence of the novel coronavirus disease, mathematical modelling has become an important tool for planning strategies to combat the pandemic by supporting decision-making and public policies, as well as allowing an assessment of the effect of different intervention scenarios. A proliferation of compartmental models was observed in the mathematical modelling community, aiming to understand and make predictions regarding the spread of COVID-19. Such approach has its own advantages and challenges: while compartmental models are suitable to simulate large populations, the underlying well-mixed population assumption might be problematic when considering non-pharmaceutical interventions (NPIs) which strongly affect the connectivity between individuals in the population. Here we propose a correction to an extended age-structured SEIR framework with dynamic transmission modelled using contact matrices for different settings in Brazil. By assuming that the mitigation strategies for COVID-19 affect the connections between different households, network percolation theory predicts that the connectivity across all households decreases drastically above a certain threshold of removed connections. We incorporated this emergent effect at population level by modulating the home contact matrices through a percolation correction function, with the few remaining parameters fitted to to hospitalisation and mortality data from the city of São Paulo. We found significant support for the model with implemented percolation effect using the Akaike Information Criteria (AIC). Besides better agreement to data, this improvement also allows for a more reliable assessment of the impact of NPIs on the epidemiological dynamics.

## 1. Introduction

Since the emergence of SARS-CoV-2, mathematical modelling has become an important tool for planning strategies to combat the pandemic by supporting decision-making and public policies [1, 2, 4, 22]. As a result, a proliferation of SEIR-like age-structured compartmental models assessing the effect of different intervention scenarios have been proposed and discussed [12, 16, 17]. As simplified representations of reality, these models come with assumptions regarding the nature of the underlying network of interactions [25]. More explicitly, most SEIR compartmental models assume that the population is homogeneously-mixed, meaning that every individual in each compartment has the same probability of contacting others, regardless of spatial distribution.

Though these underlying assumptions might be appropriate to make robust predictions for well-mixed populations, they may not be suitable in a context with significant contact network heterogeneity. Therefore, several methods have been proposed to account for contact structure heterogeneities in compartmental models [20, 26, 28]. Nevertheless, they usually consider static underlying network structures, which are only good approximations for slow epidemic spreads, in which contact rates do not change significantly over time [30, 5]. This is not the case in a pandemic such as the one caused by SARS-CoV-2, where governments were compelled to impose wide contact and movement restrictions that essentially caused a broad reduction of connectivity between individuals. The need for a more flexible and dynamic approach became evident, hence motivating the present study.

In this context, network theory provides a picture of a homogeneously mixed population as a (highly connected) regular random network of individuals (vertices) connected through possible disease transmission contacts (edges) with long-range connectivity properties [5]. During an outbreak, the disease would spread across these links. Social distancing nonpharmaceutical interventions (NPIs) could, therefore, be thought of as modulators of the strength and even persistence of such links. One of the main results of network theory is that, as contact networks become less connected, a critical transition occurs, and the network becomes fragmented into disconnected (or weakly connected) sub-networks. This phenomenon is known as percolation [7, 14], and the existence of a critical percolation threshold in the mean number of contacts is established for many kinds of networks. Across this threshold, even small changes in the number of contacts can lead to large changes in the connectivity of the network.

In compartmental models, SARS-CoV-2 mitigation strategies are modelled by altering contact rates, thus changing the force of infection. This can be done at different degrees of granularity, depending on the level of detail of contact matrices. For instance, many works [2, 21, 11] have used the categorisation employed by Prem et al. [24], which divides contacts into four settings, namely home, work, school, and other. In this way, the effectiveness of different NPIs are reflected in reductions in the contact rates for each setting, depending on the nature of the intervention – e.g. school closure reduces contact rates at school. Since the contact matrix is a linear combination of the contact matrices for each setting, with coefficients dependent on the NPIs’ coverages and efficacies, each NPI contributes linearly to reduce the infection force. By itself, this change in contacts among individuals does not affect the relationship between the mean number of contacts in the underlying network and the force of infection of the compartmental model. This is adequate if the structure of the network is not greatly affected. However, when social distancing NPIs are applied with high coverages and connections between individuals are continuously being removed, this approximation is prone to break down, and the compartmental model may no longer provide a satisfactory portrayal of the epidemiological dynamics.

To account for this change in network structure and resultant reduction in the force of infection, here we propose to modify how the effect of NPIs are modelled in compartmental models. We integrate results from percolation theory into an age-structured SEIR model by using a non-linear correction between the strength of NPIs and the resulting contact matrix, thus directly affecting the force of infection. In the following sections, we build on the previously implemented and widely used COVID-19 Modelling Consortium (CoMo) model [2], adapting its compartmentalisation to the Brazilian hospital system. Then, we present a nested model version that takes into account the loss of long-range connectivity (percolation) effect. Finally, we fit both model versions to hospitalisation and mortality data for SARS-CoV-2 during 2020 in São Paulo, Brazil, the country’s most populous city, with over 12 million inhabitants, and the first to detect COVID-19 cases. Through model comparison using the Akaike Information Criteria (AIC) [6] we find that the data strongly supports the model incorporating percolation effect.

## 2. Methods

### 2.1. Standard model

To model the epidemiological dynamics of COVID-19 in São Paulo, we apply an age-structured SEIR model with infected compartments stratified by severity of symptoms, treatment requirement and accessibility to healthcare. The main framework for this SARS-CoV-2 epidemic model was developed in collaboration with the CoMo Consortium [2] with slightly different treatment seeking compartments, an adaptation applied to better represent the Brazilian organisation of hospital beds. The progression of individuals through the infection cycle, for this version of the CoMo model, is represented by the diagram in Figure 1.

**Figure 1:**
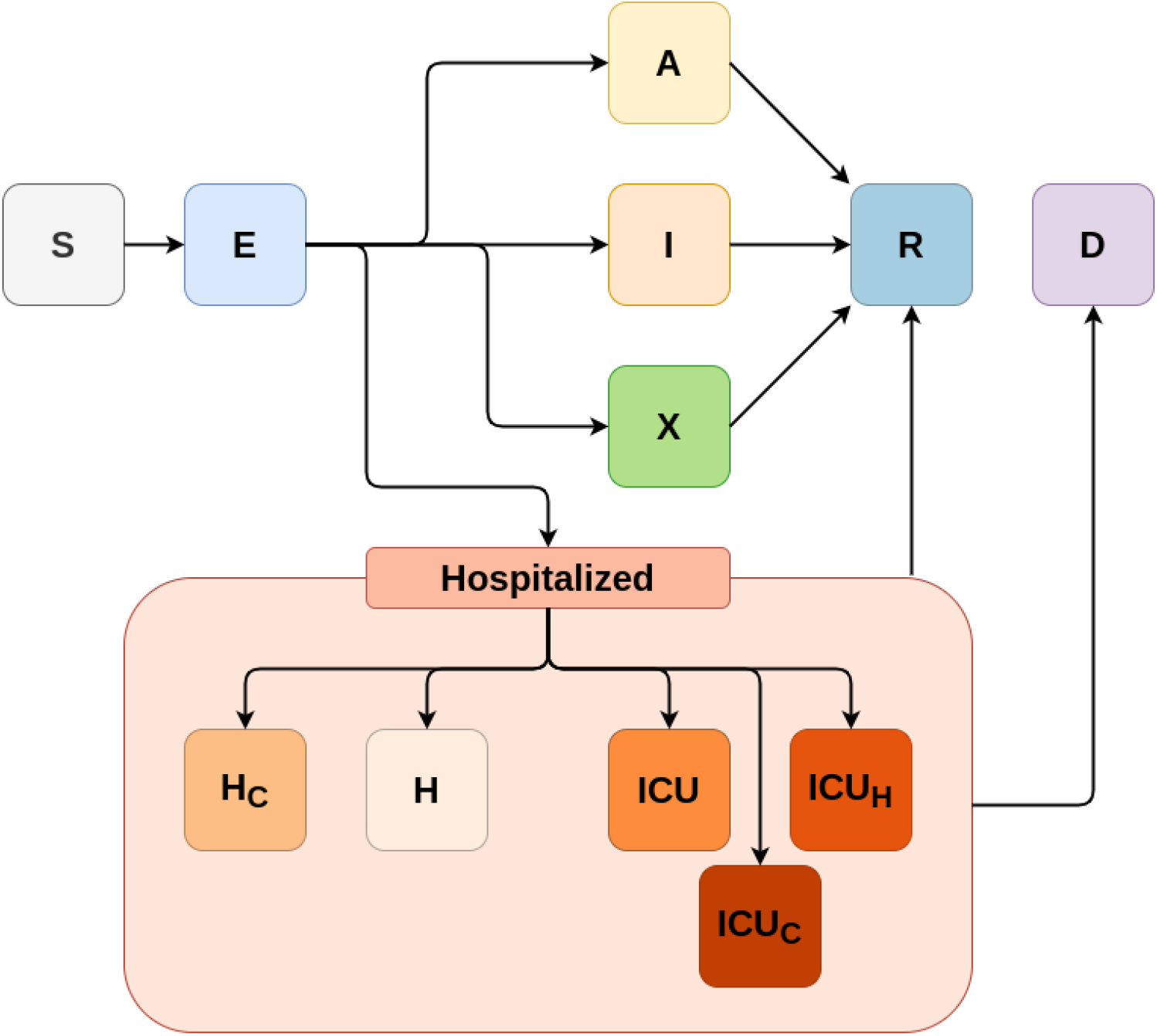
A diagram of the baseline model structure for SARS-CoV-2 in Brazil, representing the unmitigated epidemic spread scenario. The variables in the compartments represent individuals S: susceptible, E: presymptomatic infectious, A: asymptomatic infectious, I: infectious with mild symptoms, X: infectious with mild symptoms and self-isolated, H_*C*_: infectious, requiring hospital treatment but denied, H: infectious and hospitalised, ICU: infectious, receiving intensive care, ICU_*C*_: infectious, requiring intensive care but denied, ICU_*H*_ : infectious, requiring intensive care but being admitted in regular hospital bed, R: Recovered, D: Deceased. All compartments are further subdivided into 5-year age classes from 0 to 90+ years old.

Each model compartment depicted in Figure 1 is divided into 19 subcompartments, comprising all 19 age classifications from the Brazilian Institute of Geography and Statistics (IBGE) [19]. Transmission between each pair of age classes is therefore evaluated based on the contact rates between them, which are the contact matrices estimated for different settings (school, work, home, and others) in Brazil by Prem et al. [24].

We considered non-pharmaceutical intervention (NPI) scenarios including social distancing, work from home, and school closure. Here, the effect of mitigation strategies enter as linear corrections to the contact matrices, since these interventions aim to reduce contact rates or the risk of contagious at each possible contact (mask use, for example). For instance, a certain coverage on the school closure NPI would decrease contacts between all age classes in the school setting, which is modelled as a proportional decrease in contact rates in school contact matrices (see SM).

Though for school, work, and other environments such linear relationship between coverage of mitigating strategies and decrease in contact rates might be an appropriate assumption, we expect that the response should be different in the household setting. The reason for this difference is that compartmental models such as SIR-like models assume a homogeneously mixed population in all settings, which is not a good approximation for the dynamics of infection in a network of households under social distancing, as we will discuss in the next session.

### 2.2. Model with percolation

The model described in the previous session introduced a phenomenological implementation to model the effect through a proportional reduction of contact rates in the matrices. However, this approach fails to substantially decrease the transmission even when considering high coverages of NPIs. For instance, at hypothetical coverages and efficacies of 100% for school closure, work from home, and social distancing, the infection rate would still not go down immediately. Consequently, we found that data fitting was challenging when trying to simulate NPI coverages as closely as possible to the patterns seen in real life.

Further investigation suggested that the reason was the fact that, in the model, most interventions were affecting contact matrices other than the home matrices, which implied that households were simulated as fully well-mixed even under high NPI coverages. However, in reality when considering an approach of contact networks, households were becoming more isolated as other contact pathways were reduced, in such a way that the the “effective” contact rate should be much smaller. To address this issue, we draw a correspondence to percolation theory to simulate the fact that, above a certain threshold value of combined NPI adherence, inter-household contacts are much less common.

In a non-NPI scenario, we could picture the whole population as a collection of households assigned as vertices with links between households (created due to interactions at work, schools, or other places) represented by edges. This highly connected network would form one giant connected component, which could therefore be approximated as a homogeneously mixed population [5]. However, by introducing social distancing measures and increasing their coverages, we break connections in the network until long-range connectivity is lost. The critical value of number of connections (or edge density) where this transition happens is the so-called percolation threshold [14]. Above the percolation threshold of contact loss, i.e., in a high coverage social distancing scenario, we would expect to see the emergence of small household clusters that, though well connected within themselves, would be poorly connected between them.

Here we model the percolation effect on home contact matrices assuming that, while contacts outside households are kept above the percolation threshold, the effect of mitigation strategies is less apparent in home settings. On the other hand, the probability of infection of a susceptible individual drops drastically when its mean number of contacts goes below a threshold [23]. Mathematically, we correct the home contact matrix by a factor dependent on all NPIs’ coverages and efficacies, which we write as a percolation correction function, *f*_*perc*_, that has to satisfy several requirements:

- 0 ≤ *f*_*perc*_ ≤ 1;
- *f*_*perc*_ → 0 as NPIs are completely lifted; for low adherence to NPI, no connectivity loss should be noticed since different households would still be strongly connected;
- 1 − *f*_*perc*_ ≪ 1 for high effectiveness of NPIs; as connections between different households are widely severed, so *f*_*perc*_ approaches its maximum value.

A hyperbolic functional form, as follows, is proposed to model this effect:

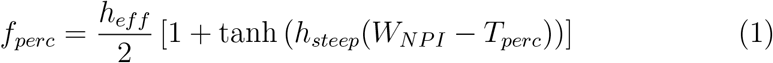

where 0 *< h*_*eff*_ *<* 1 is the maximum reduction in contacts, a limit introduced due to the percolation effect; 0 *< T*_*perc*_ *<* 1 is the percolation threshold, i.e., the fraction of connections we need to remove so that the network no longer percolates; and *h*_*steep*_ is the steepness of the percolation correction function determining how fast the network percolates: a large *h*_*steep*_ implies *f*_*perc*_ changing abruptly from 0 to *h*_*eff*_ when *W*_*NPI*_ ≈ *T*_*perc*_ (see Figure 2). Finally, *W*_*NPI*_ is defined as the weighted non-pharmaceutical interventions, combining the efficacy and coverage of each type of intervention, as well as the age and contact structure of the population they’re applied to, namely

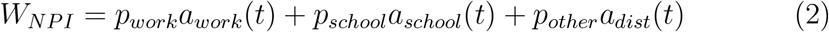

where, for instance, work adherence (*a*_*work*_) can be written as the product of the respective effectiveness (*work*_*eff*_), coverage (*work*_*cov*_) and duration of home-office strategies (translated as the *step function θ*_*work*_(*t*)):

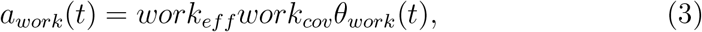

**Figure 2:**
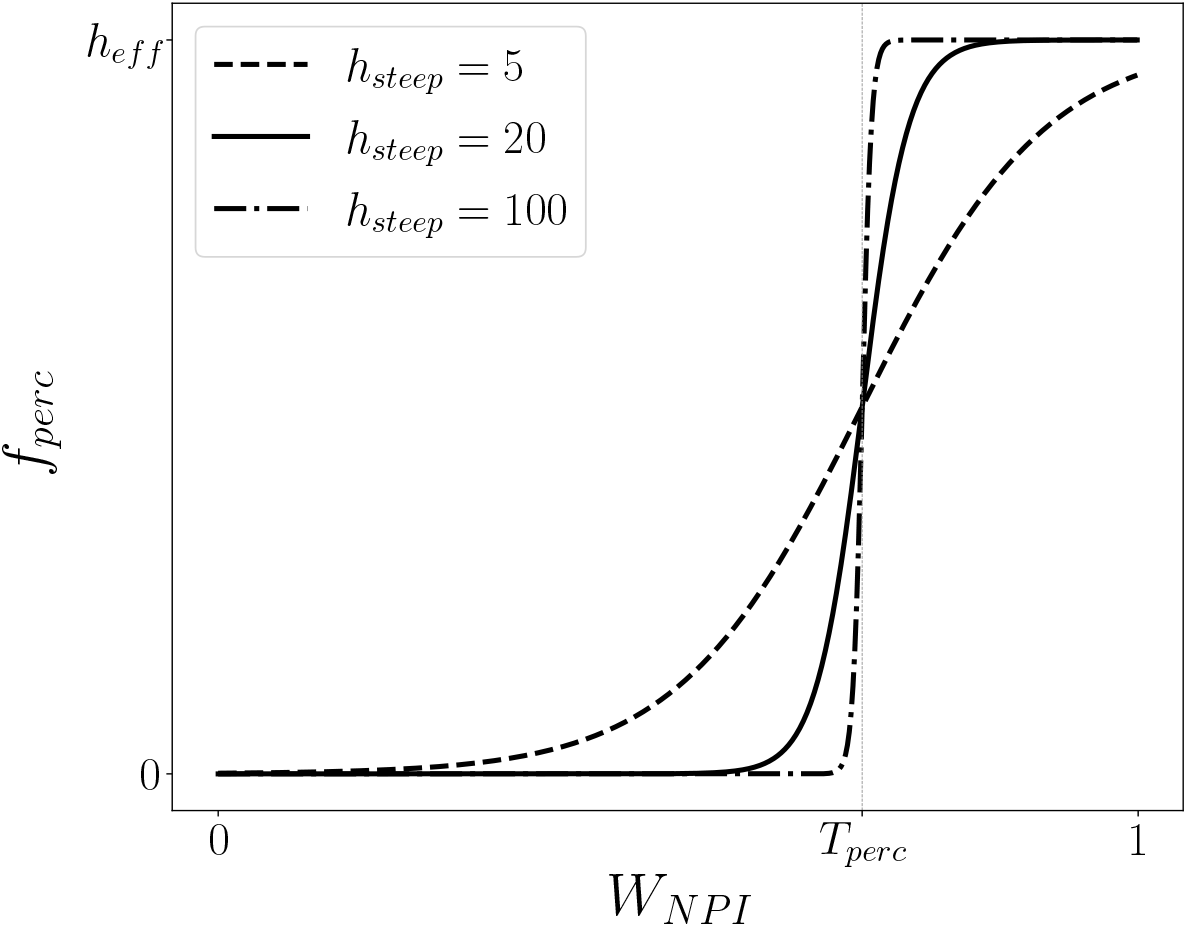
Percolation correction as a function of the combined adherence to interventions. We plot the resultant curve for different values of *h*_*steep*_, which is the steepness of the hyperbolic tangent in the definition of *f*_*perc*_ (Eq. 1). Here we used *T*_*perc*_ = 0.6 for the percolation threshold, which is indicated as the curve’s inflexion point. Note that changing its value simply dislocates the curve along the *x*-axis. For low *W*_*NP I*_, *f*_*perc*_ → 0, meaning no connectivity is lost at low adherence levels. Near *W*_*NP I*_ = *T*_*perc*_, *f*_*perc*_ grows rapidly. At the high *W*_*NP I*_ regime, where more connections are lost, *f*_*perc*_ → *h*_*eff*_ ≈ 1.

The same holds for *school* and *dist*, forming the set {*a*_*j*_(*t*)}, *j* = {*work, school, dist*}, which throughout the text will be referenced as adherences. The weights *p*_*j*_, *j* = {*work, school, other*}, are calculated as

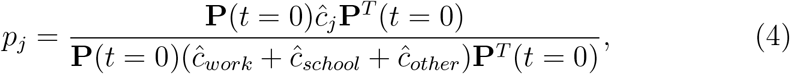

where **P**(*t* = 0) is the initial age distribution of the population and *c*_*j*_ the contact matrices in each setting.

For each NPI (*j* = {*work, school, dist*}, i.e., {work from home, school closure and social distancing}), we parametrize the respective adherence, *a*_*j*_, with efficacy and coverage parameters, here modelled as a combination of step functions allowing for changing values over time [18, 29].

Now, clearly 0 *< W*_*NPI*_ *<* 1. Note also that *W*_*NPI*_ has this specific form to consider population structure, accounting for how much each of these NPIs effectively reduce contacts in each age-class. Also note that *W*_*NPI*_ can vary according to the current implemented interventions, adding flexibility to the model.

Finally, considering all effects due to NPIs and percolation, the effective contact matrix, *ĉ*, is written as

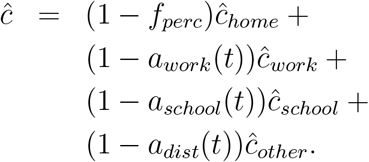

In the simulations considered here, the populations and contact matrices are obtained from publicly available data [24]; all *a*_*j*_ are estimated based on the mitigation strategies adopted by the city of São Paulo; whereas *h*_*steep*_ and *T*_*perc*_ are obtained through fitting to epidemiological data.

### 2.3. Data sources

The data used were time series of hospitalisations and deaths of Severe Acute Respiratory Illness (SARI) in São Paulo, Brazil, reported in the 2020 SIVEP-Gripe database [9]. To be sure records were already consolidated, we restricted the data to weekly aggregated data points comprising the first 23 weeks of the pandemic in the city, from 15 March to 31 August 2020.

Based on the current literature and proxy data for mobility and coverage of NPIs [18, 29, 27], we set values for all parameters with reasonable data sources or proxies (see SM for all parameter values and references). Thus only a few free parameters could not be inferred from literature and, hence, needed to be fitted to the epidemiological data. These fitted parameters were the probability of infection given a contact (*p*), start date of community transmission (*startdate*), and both *h*_*steep*_ and *T*_*perc*_ from Equation 1 for the model that takes percolation into account.

### 2.4. Fitted models

We used a Levenberg-Marquardt nonlinear least square regression fitting algorithm [13] to minimise the squared residuals, which is equivalent to maximising the likelihood under the assumption that the data points errors are normally distributed (details in SM). With that, we obtained the most probable set of values for *p, startdate, h*_*steep*_ and *T*_*perc*_. This algorithm was applied to test the two versions of the model, in three different scenarios:

- **Standard model**: derived from CoMo model [2], without including the percolation effect, which in practice meant setting *T*_*perc*_ = *h*_*steep*_ = *h*_*eff*_ = 0 and fitting only *p* and *startdate* to the data.
- **Standard model + 30% NPIs**: the model and parameters are the same as before, but we consider a much higher (30%) adherence to all NPIs. This was done to ensure that a subestimation of the effect of NPIs would not by itself favour the model with percolation – given that we already know that it always reduces the contact rates. However, notice that this leads to unrealistically high values of adherences.
- **Model with percolation**: derived from standard model, including the percolation effect, where *T*_*perc*_, *h*_*steep*_, *p* and *startdate* were fitted to the data.

Since the model with percolation introduces two extra parameters fitted to data into the standard model, we can consider them nested models and, hence, compare the quality of the fittings using the AIC.

## 3. Results

We can see the effect of implementing the percolation correction into our model by calculating the resultant basic reproduction number (*R*_0_) considering how it changes in an environment subject to NPIs. *R*_0_ is calculated using the Next Generation Matrix (NGM) method [3, Chapter 6]. In Figure 3, we see that the calculated *R*_0_ diverges among the two model versions as the combined NPI adherence approaches the fitted percolation threshold (indicated as *T*_*perc*_). Percolation implies a significantly lower *R*_0_ values for higher NPI adherence, which is consistent with the steep decrease in NPI effect near the percolation threshold modelled by the percolation correction function (Eq. 1).

**Figure 3:**
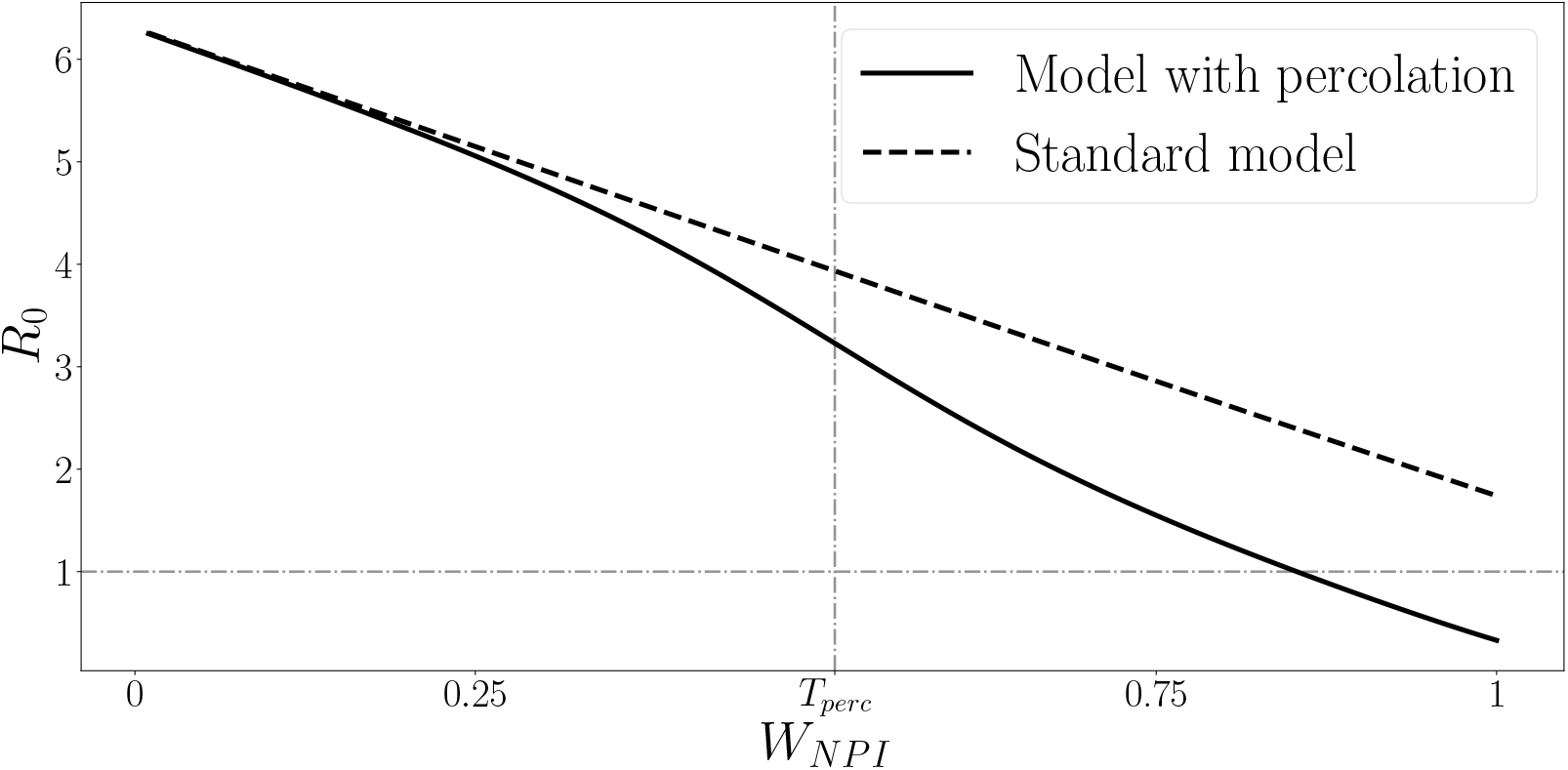
Resultant epidemic basic reproduction number (*R*_0_) as a function of the combined NPI adherence for both model versions. Note the steep diverence near the percolation threshold (*T*_*perc*_ = 0.514, as resultant from fitting the model with percolation to data).

After implementing both model versions with equivalent fixed parameter sets, and also with increased NPIs in one scenario (see complete table of parameters in the SM), we fitted the free parameters (among *T*_*perc*_, *h*_*steep*_, *p* and *startdate*) in each model to weekly new cases and new deaths from 2020 SARI data for São Paulo.

Figure 4 shows the resulting curves for all fitted models, with confidence intervals assuming that fitted parameters follow a multivariate normal distribution. These confidence intervals are estimated by bootstrapping the covariance matrix from the Levenberg-Marquardt algorithm. The comparison between the two standard model curves shows that increasing intervention coverages (even to the point of assuming unrealistic values) can modulate the epidemic curve to a certain extent, but still does not result in a good fit to data.

**Figure 4:**
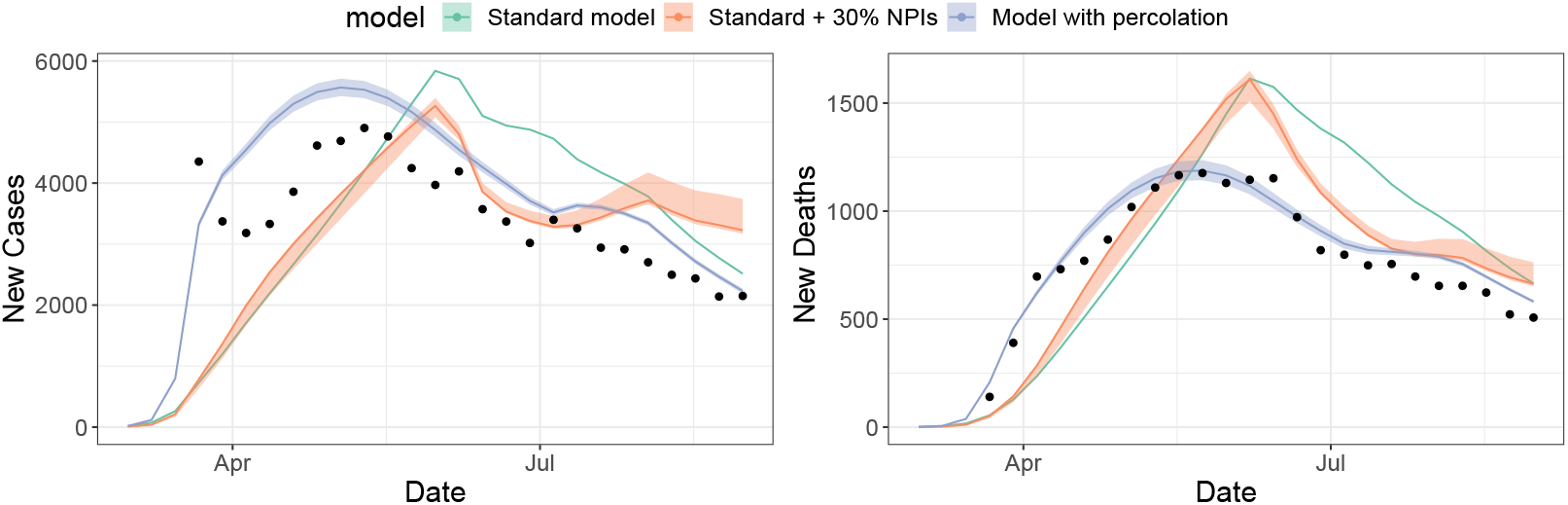
Results from simultaneous model fitting to SARI new cases and new deaths data, for each of the proposed model versions: the model with percolation (blue) and the standard model (green), using the same parameter sets, apart from the fitted parameters (*p, startdate, T*_*perc*_, *h*_*steep*_), and the standard model version with 30% more coverage of all NPIs (red) (also fitted to data); dots represent data points from SIVEP-Gripe database (15 March to 31 August 2020) [9]; and shades represent bootstrapped confidence intervals based on parameter uncertainty from the fitting (too thin to see for the green curve).

The resulting parameters and the computed AIC for each model version are shown in Table 1. Even though in Figure 4 the curves for the *standard model + 30% NPI* and the *model with percolation* might look somewhat close, the AIC clearly shows a much higher level of empirical support for the model version with percolation.

**Table 1:** Fitted parameter values and AIC evaluated for each of the model versions, as they were fitted to the same dataset of COVID-19 weekly cases and deaths for São Paulo, which consisted of 23 data points. ΔAIC is the difference between the AIC for each model and the minimum overall AIC.

| Model | $p$ | $startdate$ | $h_{steep}$ | $T_{perc}$ | AIC | $\Delta AIC$ |
| --- | --- | --- | --- | --- | --- | --- |
| Standard | 0.0294 | 2020-01-15 | - | - | -756 | 191 |
| Standard + 30% NPI | 0.0401 | 2020-01-30 | - | - | -793 | 154 |
| Percolation | 0.0461 | 2020-01-30 | 4.83 | 0.516 | -947 | 0 |

## 4. Discussion

Modelling the ongoing COVID-19 pandemic represented an unprecedented challenge for modellers, as we tried to make urgent recommendations and predictions with scarce information about the virus’s underlying biology, its main spread mechanisms, and severity and fatality rates [10, 8]. Furthermore, we lacked individual level data on mobility patterns to accurately understand the dynamic effect of social distancing NPIs on individual behaviour and contact patterns.

Although previous works considered heterogeneous contact structures in epidemiological models [20, 26, 28], and many others accounted for NPIs’ effects on transmissibility [12, 16, 17, 1, 2, 4, 22], few to none made the connection between both phenomena. Strictly speaking, NPIs can affect the underlying network structure over time due to large scale social distancing interventions. In this context, we identified a demand for more flexible and dynamic modelling approaches taking into account the non-linear phenomena emerging from social distancing NPIs.

In the case of the COVID-19 pandemic, governments enforced wide movement and contact restrictions, essentially removing connections between individuals. To take into account the structural heterogeneities introduced by these social distancing measures on a population’s contact network, we proposed a phenomenological correction to compartmental models that only depends on quantities already computed when modelling NPIs, without the addition of an excessive number of parameters which could lead to model overfitting. Using the Akaike Information Criterion (AIC) we found ΔAIC = 191 between the model with percolation and the standard model (Table 1), which is orders of magnitude higher than the conventional threshold of Δ*AIC* ≥ 2 to consider significant support for one model [15], that is, the model with percolation correction had very strong empirical support with the data analysed.

One alternative explanation could be that it might still be possible to adjust other model parameters to obtain a better fit to data in the standard model by adjusting NPIs’ coverages. However, with further exploration of the parameter space, even considering less realistic parameter sets (e.g. tampering with NPIs’ coverage parameters), we could not obtain a good fit to data. For instance, a 30% increase on NPI coverage on the standard model (red curve in Figure 4) resulted in ΔAIC = 154. Though it represented a better fitting compared to the standard model version, it was not as good as the one obtained with the version including the percolation.

These results highlight the importance when using compartmental models of implementing this non-linear response to social distancing NPIs in order to obtain a better representation of its effect on a large population.

## 5. Conclusion

The correction to an age-structured SEIR compartmental model described here, though motivated by the need to better represent the COVID-19 spread dynamics, was inspired by network theory and general individual level considerations that affected a population’s underlying network structure. We implemented the effect of individual behaviour changes on a population’s macroscopic dynamics without drastically increasing the number of fitted parameters in our compartmental model.

The usefulness of incorporating this fragmentation process into our SARS-CoV-2 spread compartmental model was evident, but note that its flexibility also permits it to be applied when modelling the transmission dynamics of other communicable diseases under similar high NPIs’ coverage regimes.

Therefore, our framework may be applied to any homogeneously-mixing compartmental models trying to represent the dynamics of a population suffering drastic changes in its connectivity patterns during an epidemic. This result contributes towards a more accurate epidemic modelling, potentially implying in better control and prevention policy recommendations at public health level.

## Supporting information

Model equations; parameterization; contact matrices and force of infection; hospital burden; fitting procedure

## Data Availability

The algorithm developed in this work is available in our GitHub Repository.

https://github.com/francocarol/covid_perc

## Authors’ contributions

Conceptualization: CF, LSF, VS, SP, RMC and RAK. Formal analysis and Software: CF, LSF, MEB, SP and RMC. Writing - Original Draft, Review & Editing: CF, LS, VS, SP and RMC. Data Curation: PIP. Supervision and Writing - Review & Editing: RA, LW and RAK.

## Declaration of Competing Interest

We have no competing interests.

## Acknowledgements

We thank all members of Observatório COVID-19 BR and CoMo Consortium for the collaborative work. The authors also thank the research funding agencies: São Paulo Research Foundation (FAPESP) - Brazil (grant number: 2019/26310-2 and 2017/26770-8 to CF, 2018/24037-4 to SP, 2018/23984-0 to VS and contract number: 2016/01343-7 to RAK), Coordenação de Aperfeiçoamento de Pessoal de Nível Superior – Brazil (CAPES) (Finance Code 001 to LSF) and the Brazilian National Council for Scientific and Technological Development (CNPq) (grant number: 315854/2020-0 to MEB, 313055/2020-3 to PIP and 311832/2017-2 to RAK). RA is funded by the Bill and Melinda Gates Foundation (OPP1193472). LW is funded by the Li Ka Shing Foundation. The CoMo Consortium has support from the Oxford University COVID-19 Research Response Fund (ref: 0009280).

## Appendix A

R codes (https://github.com/francocarol/covid_perc) and supplementary material are provided.

## Notes

### Competing Interest Statement

The authors have declared no competing interest.

### Author Declarations

Ethics approval was not necessary because this study analysed only publicly available data, not including identifiable information.

## References

[1] D. Adam. Special report: The simulations driving the world’s response to COVID-19. Nature, 580(7802):316–319, 2020.

[2] R. Aguas, L. White, N. Hupert, R. Shretta, W. Pan-Ngum, O. Celhay, A. Moldokmatova, F. Arifi, A. Mirzazadeh, H. Sharifi, K. Adib, M. N. Sahak, C. Franco, and R. Coutinho. Modelling the COVID-19 pandemic in context: an international participatory approach. BMJ Global Health, 5(12), 2020. doi:10.1136/bmjgh-2020-003126. URL https://gh.bmj.com/content/5/12/e003126.

[3] L. J. Allen, F. Brauer, P. Van den Driessche, and J. Wu. Mathematical epidemiology, volume 1945. Springer, 2008.

[4] A. Anirudh. Mathematical modeling and the transmission dynamics in predicting the Covid-19 - What next in combating the pandemic. Infectious Disease Modelling, 5:366–374, 2020.

[5] S. Bansal, B. T. Grenfell, and L. A. Meyers. When individual behaviour matters: homogeneous and network models in epidemiology. Journal of the Royal Society Interface, 4(16):4879—-891, October 2007. ISSN 1742-5662. doi:10.1098/rsif.2007.1100. URL https://royalsocietypublishing.org/doi/10.1098/rsif.2007.1100.

[6] K. P. Burnham and D. R. Anderson. Model Selection and multi-model inference: A practical information-theoretic approach. Springer New York, 2013.

[7] Y. Chen, G. Paul, R. Cohen, S. Havlin, S. P. Borgatti, F. Liljeros, and H. Eugene Stanley. Percolation theory and fragmentation measures in social networks. Physica A: Statistical Mechanics and its Applications, 378(1):11–19, 2007. ISSN 0378-4371. doi:https://doi.org/10.1016/j.physa.2006.11.074. URL https://www.sciencedirect.com/science/article/pii/S0378437106012611.

[8] D. Cyranoski. Profile of a killer: the complex biology powering the coronavirus pandemic. Nature, 581(780 6):22–26, May 2020. doi:10.1038/d41586-020-01315-73.

[9] Datasus. Srag 2020 - banco de dados de síndrome respiratória aguda grave - incluindo dados da covid-19, 2020. URL https://opendatasus.saude.gov.br/dataset/bd-srag-2020.

[10] G. Davey Smith, M. Blastland, and M. Munafò. Covid-19’s known unknowns. BMJ, 371, 2020. doi:10.1136/bmj.m3979. URL https://www.bmj.com/content/371/bmj.m3979.

[11] N. G. Davies, P. Klepac, Y. Liu, K. P. A. M. Jit, C. C.-. working group, and R. M. Eggo. Age-dependent effects in the transmission and control of COVID-19 epidemics. Nature Medicine, 26:1205–1211, 2020. doi:10.1038/s41591-020-0962-9.

[12] N. G. Davies, A. J. Kucharski, R. M. Eggo, A. Gimma, W. J. Edmunds, T. Jombart, K. O’Reilly, A. Endo, J. Hellewell, E. S. Nightingale, et al. Effects of non-pharmaceutical interventions on covid-19 cases, deaths, and demand for hospital services in the UK: a modelling study. The Lancet Public Health, 5(7):e375–e385, 2020.

[13] T. V. Elzhov, K. M. Mullen, A.-N. Spiess, and B. Bolker. minpack.lm: R Interface to the Levenberg-Marquardt Nonlinear Least-Squares Algorithm Found in MINPACK, Plus Support for Bounds, 2016. URL https://CRAN.R-project.org/package=minpack.lm. R package version 1.2-1.

[14] J. W. Essam. Percolation theory. Reports on Progress in Physics, 43 (7):833–912, July 1980. doi:10.1088/0034-4885/43/7/001. URL https://doi.org/10.1088/0034-4885/43/7/001.

[15] F. J. Fabozzi, S. M. Focardi, S. T. Rachev, and B. G. Arshanapalli. Appendix E: Model Selection Criterion: AIC and BIC, pages 399–403. John Wiley & Sons, Ltd, 2014. ISBN 9781118856406. doi:https://doi.org/10.1002/9781118856406.app5. URL https://onlinelibrary.wiley.com/doi/abs/10.1002/9781118856406.app5.

[16] N. Ferguson, D. Laydon, G. Nedjati Gilani, N. Imai, K. Ainslie, M. Baguelin, S. Bhatia, A. Boonyasiri, Z. Cucunuba Perez, G. Cuomo-Dannenburg, et al. Report 9: Impact of non-pharmaceutical interventions (NPIs) to reduce covid19 mortality and healthcare demand. Technical report, Imperial College London, 2020.

[17] S. Flaxman, S. Mishra, A. Gandy, H. J. T. Unwin, T. A. Mellan, H. Coupland, C. Whittaker, H. Zhu, T. Berah, J. W. Eaton, et al. Estimating the effects of non-pharmaceutical interventions on COVID-19 in europe. Nature, 584(7820):257–261, 2020.

[18] Google. Covid-19 mobility reports. Technical report, Google, 2020. URL https://www.google.com/covid19/mobility/.

[19] IBGE. Metodologia do censo demográfico 2010. IBGE, Rio de Janeiro, 2016. ISBN 9788524043628. URL https://biblioteca.ibge.gov.br/index.php/biblioteca-catalogo?view=detalhes&id=296501.

[20] W. min Liu, H. W. Hethcote, and S. A. Levin. Dynamical behavior of epidemiological models with nonlinear incidence rates. Journal of Mathematical Biology, 25(4):359–380, Sept. 1987. doi:10.1007/bf00277162. URL https://doi.org/10.1007/bf00277162.

[21] N. B. Noll, I. Aksamentov, V. Druelle, A. Badenhorst, B. Ronzani, G. Jefferies, J. Albert, and R. A. Neher. Covid-19 scenarios: an interactive tool to explore the spread and associated morbidity and mortality of sars-cov-2. medRxiv, 2020. doi:10.1101/2020.05.05.20091363. URL https://www.medrxiv.org/content/early/2020/05/12/2020.05.05.20091363.

[22] J. Panovska-Griffiths, C. Kerr, W. Waites, and R. Stuart. Mathematical modeling as a tool for policy decision making: Applications to the COVID-19 pandemic. Elsevier, 2021.

[23] R. Pastor-Satorras, C. Castellano, P. Van Mieghem, and A. Vespignani. Epidemic processes in complex networks. Rev. Mod. Phys., 87:925–979, Aug 2015. doi:10.1103/RevModPhys.87.925. URL https://link.aps.org/doi/10.1103/RevModPhys.87.925.

[24] K. Prem, K. van Zandvoort, P. Klepac, R. M. Eggo, N. G. Davies, A. R. Cook, and M. Jit. Projecting contact matrices in 177 geographical regions: an update and comparison with empirical data for the COVID-19 era. medRxiv, 2020. doi:10.1101/2020.07.22.20159772. URL https://www.medrxiv.org/content/early/2020/07/28/2020.07.22.20159772.

[25] H. Rahmandad and J. Sterman. Heterogeneity and network structure in the dynamics of diffusion: Comparing agent-based and differential equation models. Management Science, 54(5):998–1014, May 2008. doi:10.1287/mnsc.1070.0787. URL https://doi.org/10.1287/mnsc.1070.0787.

[26] M. Roy and M. Pascual. On representing network heterogeneities in the incidence rate of simple epidemic models. Ecological Complexity, 3(1):80–90, Mar. 2006. doi:10.1016/j.ecocom.2005.09.001. URL https://doi.org/10.1016/j.ecocom.2005.09.001.

[27] Secretaria Municipal de Mobilidade e Transportes, Cidade de São Paulo. Passageiros transportados - 2020, 2020. URL https://www.prefeitura.sp.gov.br/cidade/secretarias/transportes/institucional/sptrans/acessoainformacao/agenda/index.php?p=292723.

[28] P. D. Stroud, S. J. Sydoriak, J. M. Riese, J. P. Smith, S. M. Mniszewski, and P. R. Romero. Semi-empirical power-law scaling of new infection rate to model epidemic dynamics with inhomogeneous mixing. Mathematical Biosciences, 203(2):301–318, Oct. 2006. doi:10.1016/j.mbs.2006.01.007. URL https://doi.org/10.1016/j.mbs.2006.01.007.

[29] C. M. Toscano, A. F. R. Lima, L. L. S. Silva, P. F. Razia, L. F. A. Pavão, D. A. Polli, R. F. Moraes, and M. A. Cavalcanti. Medidas de distanciamento social e evolução da covid-19 no brasil, 2020. URL https://medidas-covidbr-iptsp.shinyapps.io/painel/.

[30] E. Volz and L. A. Meyers. Susceptible&#x2013;infected&#x2013;recovered epidemics in dynamic contact networks. Proceedings of the Royal Society B: Biological Sciences, 274(1628):2925–2934, 2007. doi:10.1098/rspb.2007.1159. URL https://royalsocietypublishing.org/doi/abs/10.1098/rspb.2007.1159.

