## Supplementary material for "Percolation across households in mechanistic models of non-pharmaceutical interventions in SARS-CoV-2 disease dynamics": Model equations; parameterization; contact matrices and force of infection; hospital burden; fitting procedure

[1]*Institute of Theoretical Physics, São Paulo State University, São Paulo, Brazil*
[2]*Big Data Institute, Li Ka Shing Centre for Health Information and Discovery, Nuffield Department of Medicine, University of Oxford, Oxford, UK*
[3]*Observatório Covid-19 BR*
[4]*Department of Ecology and Evolution, University of Lausanne, Lausanne, Switzerland*
[5]*Instituto de Biociências, Universidade de São Paulo, São Paulo, Brazil*
[6]*Nuffield Department of Medicine, University of Oxford Centre for Tropical Medicine and Global Health, Oxford, Oxfordshire, UK*
[7]*Centro de Matemática, Computação e Cognição - Universidade Federal do ABC, Santo André, Brazil*

### I. Introduction

This model framework was first introduced in Aguas et al. (1). The code is available at https://github.com/francocarol/covid_perc. In Section II we introduce our modifications for the CoMo model structure (1) to account for particularities in the Brazilian health system. Nevertheless, modifications to better reflect other localities should be easily included in the model. Section II-A describes the equations, along with the explanation and sources of the parameters used. Section II-B describes how non-pharmaceutical interventions work in the model. Section II-C thoroughly describes our modifications to model hospital burden in Brazil. Section III lists the interventions used in the main paper. Finally, Section IV shows the procedure used to fit the model to data.

### II. Model structure

#### A. Model equations

The model consists in an expanded SEIR model to account for asymptomatic individuals and detailed structure of the Brazilian health system. We write

$$
\begin{aligned}
\frac{d\mathbf{S}}{dt} &= -\lambda\mathbf{S} + \omega\mathbf{R} + \mathbf{A}\cdot\mathbf{S} + \mu_b - \mu_d\mathbf{S} \\
\frac{d\mathbf{E}}{dt} &= \lambda\mathbf{S} - \gamma\mathbf{E} + \mathbf{A}\cdot\mathbf{E} - \mu_d\mathbf{E} \\
\frac{d\mathbf{I}}{dt} &= \gamma(1-P_{clin})(1-IHR)\mathbf{E} - \nu_i\mathbf{I} + \mathbf{A}\cdot\mathbf{I} - \mu_d\mathbf{I} \\
\frac{d\mathbf{CL}}{dt} &= \gamma P_{clin}(1-P_{selfis})(1-IHR)\mathbf{E} - \nu_i\mathbf{CL} + \mathbf{A}\cdot\mathbf{CL} - \mu_d\mathbf{CL} \\
\frac{d\mathbf{X}}{dt} &= \gamma P_{selfis}P_{clin}(1-IHR)\mathbf{E} - \nu_i\mathbf{X} + \mathbf{A}\cdot\mathbf{X} - \mu_d\mathbf{X} \\
\frac{d\mathbf{H}}{dt} &= \gamma IHR(1-P_{icu})(1-H_c)\mathbf{E} - \nu_s\mathbf{H} + \mathbf{A}\cdot\mathbf{H} - \mu_d\mathbf{H} \\
\frac{d\mathbf{HC}}{dt} &= \gamma IHR(1-P_{icu})H_c\mathbf{E} - \nu_{sc}\mathbf{HC} + \mathbf{A}\cdot\mathbf{HC} - \mu_d\mathbf{HC} \\
\frac{d\mathbf{ICU}}{dt} &= \gamma IHRP_{icu}(1-ICU_c)\mathbf{E} - \nu_{icu}\mathbf{ICU} + \mathbf{A}\cdot\mathbf{ICU} - \mu_d\mathbf{ICU} \\
\frac{d\mathbf{ICUH}}{dt} &= \gamma IHRP_{icu}ICU_c(1-ICUH_c)E - \nu_{icuh}\mathbf{ICUH} + \mathbf{A}\cdot\mathbf{ICUH} - \mu_d\mathbf{ICUH} \\
\frac{d\mathbf{ICUC}}{dt} &= \gamma IHRP_{icu}ICU_cICUH_c\mathbf{E} - \nu_{icuc}\mathbf{ICUC} + \mathbf{A}\cdot\mathbf{ICUC} - \mu_d\mathbf{ICUC} \\
\frac{d\mathbf{R}}{dt} &= \nu_i\mathbf{I} - \omega\mathbf{R} + \nu_i\mathbf{X} + \nu_i\mathbf{CL} + \mathbf{A}\cdot\mathbf{R} - \mu_d\mathbf{R} + \nu_s(1-P_dI_{fr})\mathbf{H} \\
&+ \nu_{icu}(1-P_{dicu}IHFR)\mathbf{ICU} + \nu_{icuc}(1-P_{dicuc}IHFR)\mathbf{ICUC} + \nu_{sc}(1-P_{dhc}IHFR)\mathbf{HC} \\
&+ \nu_{icuh}(1-P_{dicuh}IHFR)\mathbf{ICUH} + \nu_{icuc}(1-P_{dicuc}IHFR)\mathbf{ICUC}
\end{aligned}
$$

| code and equations | description |
|---|---|
| **S** | Susceptible population |
| **E** | Infected and presymptomatic population |
| **I** | Infected population, asymptomatic and not isolated |
| **CL** | Infected population, mildly symptomatic and not isolated |
| **X** | Infected population, mildly symptomatic and self-isolated at home |
| **H** | Infected population, hospitalized in simple bed. |
| **HC** | Infected population that require hospital treatment but but are denied, due to healthcare system overload |
| **ICU** | Infected population, hospitalised in Intensive Care Units (ICU). |
| **ICUH** | Infected population that require ICU but are hospitalised in simple beds, due to unavailability in ICU beds. |
| **ICUC** | Infected population that require ICU but are denied both an ICU or hospital simple bed, due to healthcare system overload. |
| **R** | Recovered population |
| **C** | Cumulative reported cases |
| $\mathbf{C_M}$ | Cumulative death cases |
| $\mathbf{C_{MC}}$ | Cumulative death cases of critical patients, i.e., those who hospitalization was denied. |

**TABLE I:** List of model variables in equations on supplementary material and in the code. Variables written in the main text may be different for readability, here, we stick to the nomenclature used throughout the code to help reproducibility.

$$\frac{d\mathbf{C}}{dt} = r\gamma(1-IHR)(1-P_{clin})\mathbf{E} + r_c\gamma(1-IHR)P_{clin}\mathbf{E} + r_h\gamma IHR\mathbf{E}$$

$$\frac{d\mathbf{CM}}{dt} = \nu_s P_{dh} IHFR\mathbf{H} + \nu_{sc}P_{dhc}IHFR\mathbf{HC} + \nu_{icu}P_{dicu}IHFR\mathbf{ICU} + \nu_{icuc}P_{dicuc}IHFR\mathbf{ICUC}$$
$$+ \nu_{icuh}P_{dicuh}IHFR\mathbf{ICUH} + \mu_d(\mathbf{H}+\mathbf{HC}+\mathbf{ICU}+\mathbf{ICUC}+\mathbf{ICUH}+\mathbf{CL}+\mathbf{X})$$

$$\frac{d\mathbf{CMC}}{dt} = \nu_{sc}P_{dhc}IHFR\mathbf{HC} + \nu_{icuc}P_{dicuc}IHFR\mathbf{ICUC}$$
$$+ \nu_{icuh}P_{dicuh}IHFR\mathbf{ICUH} + \mu_d(\mathbf{HC}+\mathbf{ICUC})$$

where each of the dynamic variables (corresponding to the compartments shown in Table I) is further subdivided in 19 age classes consisting of 5 years age bins (0-4,5-9, up to 90+). Thereby, each of the parameters written in the model, aside from **A** (ageing matrix), should be thought of as diagonal matrices containing parameter values corresponding to each age class. Take, as an example, the natural mortality rate, given by

$$\hat{\mu_d} = \text{diag}(\mu_{d1}, \mu_{d2}, ..., \mu_{dD}) = \text{diag}(\vec{\mu_d}).$$

Note that, in the system of equations presented above, we drop the hats/bolds from all diagonal matrices to avoid an overloaded notation, but choose to keep them in all variables. Thus, each of them actually represents $D = 19$ different ODEs, and therefore the number of equations is $D$ multiplied by the number of compartments. A description of each parameter from the model is available at table II.

#### *B. Contact matrices and the force of infection*

The model has over 300 equations, but the main mechanisms regarding infection and NPI effects are encoded in its force of infection, $\lambda$, which is describe in this subsection. Basically, a Susceptible individual in the $n$-th age class, written as $S_n$, can have contact with any of the infected groups of all age classes, so we would have

$$\frac{dS_n}{dt} \propto -S_n \sum_j c_{n,j}(a_j^I I_j + a_j^E E_j + a_j^{CL} CL_j + ...) \tag{1}$$

where $c_{n,j}$ measures the contact strength between people of $n$-th and $j$-th age classes, forming the contact matrix $\hat{c}$, of dimension $D \times D$. The $a_j^I, a_j^E, a_j^{CL}, ...$ measures how infectious these different model compartments are. For example, asymptomatic people, **I**, may be more infectious than the symptomatic ones, **CL**, since they may not be isolating themselves, given they are unaware of their infectious state. The NPIs are considered as modifications on both $\hat{c}$, reducing contacts between people, and also on the different $a_j^I, a_j^{CL}, ..$, accounting for behavioural aspects, such as increased hand hygiene.

Given the definitions above, we are finally able to breakdown the general structure of $\hat{c}$. It is mainly composed of 4 matrices, $\hat{c}_{home}$ - which measures the amount of contacts of people at home, $\hat{c}_{work}$ - for contacts at work, $\hat{c}_{school}$ - for contacts at school and $\hat{c}_{other}$ - for other kinds of human interactions, such as going to restaurants, movies and churches. Therefore, in absence of any NPIs, the resultant contacts matrix would be simply

$$\hat{c} = \hat{c}_{home} + \hat{c}_{work} + \hat{c}_{school} + \hat{c}_{other}$$

But as NPIs are inserted, the contact matrix is modified. Suppose the simple case of home-office policies, that is, people should work at home for a period of $work_{dur}$ weeks. A fraction $work_{cov}$ of the population is able to adhere to such policies, and they have an effectiveness $work_{eff}$ in reducing this kind of contact, then we have that the contact matrix becomes

$$\hat{c} = (1 - f_{perc})\hat{c}_{home} + (1 - work_{cov}work_{eff}\theta_{work}(t))\hat{c}_{work} + \hat{c}_{school} + \hat{c}_{other}$$

| Code | Equation | Description | Value | Source |
|---|---|---|---|---|
| lam | $\lambda$ | force of infection | Variable | Eq. (5) |
| mort | $\mu_d$ | natural mortality ($days^{-1}$) | Age dependent | IBGE (7) |
| ageing | $\mathbf{A}$ | speed of population ageing ($days^{-1}$) | - | - |
| birth | $\mu_b$ | birth rate ($days^{-1}$) | - | IBGE (8) |
| gamma | $\gamma$ | Inverse of incubation period ($days^{-1}$) | $1/5.8$ | Wei et al. (13) |
| ihr | IHR | Infection hospitalisation rate | Age dependent | Salje et al. (10) |
| omega | $\omega$ | Rate of which recovered people become susceptible again ($days^{-1}$) | 0 | Assumed |
| rho | $\rho$ | Relative infectiousness of presymptomatic individuals | 0.105 | Wei et al. (12) |
| rhos | $\rho_s$ | Relative infectiousness of hospitalised individuals (reduced due to hospitalisation) | 0.10 | Assumed |
| pclin | $P_{clin}$ | Proportion of symptomatic individuals | 0.30 (0-19)<br>0.56 (20-59)<br>0.69 (60+) | SMSSP<br>Sun et al. (11)<br>Sun et al. (11) |
| selfis | $P_{selfis}$ | Proportion of symptomatic individuals who self-isolate | Variable | Section II-B |
| prob_icu | $P_{icu}$ | Proportion of hospitalised individuals who need ICU beds | Age dependent | SIVEP |
| critH | $H_c$ | Proportion of hospitalised individuals who have not received attendance | Variable | Section II-C |
| critICU | $ICU_c$ | Proportion of hospitalised individuals who need ICU beds and have not received one | Variable | Section II-C |
| critICUH | $ICU_h$ | Proportion of hospitalised individuals who need ICU beds and have not received one and also not have received simple beds | Variable | Section II-C |
| nui | $\nu_i$ | Recovery rate of mild symptomatic/asymptomatic individuals ($days^{-1}$) | 1/9 | Cevik et al. (3) |
| nus | $\nu_s$ | Recovery/death rate of hospitalised individuals ($days^{-1}$) | 1/8.3 | SIVEP |
| nusc | $\nu_{sc}$ | Recovery/death rate of hospitalised individuals who have not received attendance ($days^{-1}$) | 1/11 | Assumed |
| nu_icu | $\nu_{icu}$ | Recovery/death rate of hospitalised individuals in ICU beds ($days^{-1}$) | 1/14.7 | SIVEP |
| nu_icuh | $\nu_{icuh}$ | Recovery/death rate of hospitalised individuals who need ICU beds but received simple beds ($days^{-1}$) | 1/11 | Assumed |
| nu_icuc | $\nu_{icuc}$ | Recovery/death rate of hospitalised individuals who need ICU beds and have not received attendance ($days^{-1}$) | 1/11 | Assumed |
| ifr | IHFR | In hospital fatality rate | Age dependent | Portella et al. (9) |
| pdeath_h | $P_d$ | Maximum probability of death for a hospitalised infection requiring common bed | 45.9 | SIVEP |
| pdeath_icu | $P_{dicu}$ | Maximum probability of death for a hospitalised infection requiring ICU | 69 | SIVEP |
| pdeath_hc | $P_{dhc}$ | Maximum probability of death for a hospitalised infection requiring common bed but not receiving attendance | 80 | Assumed |
| pdeath_icuh | $P_{dicuh}$ | Maximum probability of death for a hospitalised infection requiring ICU but receiving common bed attendance | 97 | Assumed |
| pdeath_icuc | $P_{dicuc}$ | Maximum probability of death for a hospitalised infection requiring ICU but not receiving attendance | 99 | Assumed |
| report | r | Report rate of asymptomatic cases | 0.00 | Assumed |
| reportc | $r_c$ | Report rate of symptomatic cases | 0.01 | Assumed |
| reporth | $r_h$ | Report rate of hospitalized cases | 0.95 | Assumed |

**TABLE II:** List of model parameters in equations on supplementary material and in the code. These variables are restricted to epidemiological variables (not the NPI-related ones).

Here, $f_{perc}$ is as defined as in section 2.2 of the main paper, whilst $\theta_{work}(t)$ is a function that measures if either home office policies are being applied or not, and is usually a step function, being 1 during the period $work_{dur}$, defined by starting and finishing dates of such policies, and 0 out of said period. We could apply this same mathematical form for other interventions, such as school closing, commerce and restaurants functioning in reduced periods or not functioning at all, and many other NPIs we have seen being tried out in order to contain virus spread. We would get

$$\begin{aligned}\hat{c} \;=\; & (1 - f_{perc})\hat{c}_{home} + \\ & (1 - work_{cov} work_{eff} \theta_{work}(t))\hat{c}_{work} + \\ & (1 - school_{cov} school_{eff} \theta_{school}(t))\hat{c}_{school} + \\ & (1 - dist_{cov} dist_{eff} \theta_{dist}(t))\hat{c}_{other}\end{aligned}$$

With this final form of contact matrix we can add yet another possible NPI, cocooning the elderly. That means they are more isolated and protected, since they are one of the most vulnerable to COVID-19 death. When cocooning is applied, the contacts of people above a certain age, let's say $age_{cocoon}$, are reduced in $cocoon_{cov} cocoon_{eff}$, implying that $\hat{c}$ values from the $D^{\dagger}$ to $D$ lines and rows must be reduced. Here, $D^{\dagger}$ is the index from which cocooning starts, for example, if cocooning is applied in people over 65 years old, then $D^{\dagger} = 13$. Defining $\eta = 1 - cocoon_{cov} cocoon_{eff} \theta_{cocoon}(t)$ we write

$$\hat{g}_{D^{\dagger}}(\eta) = \mathrm{diag}(\vec{1}_{D^{\dagger}}, \eta \vec{1}_{D-D^{\dagger}}) \tag{2}$$

and with it, make the final contacts matrix

$$\bar{c} \;=\; \hat{g}_{D^{\dagger}}(\eta) \hat{c} \hat{g}_{D^{\dagger}}(\eta) \tag{3}$$

Note that

$$\hat{g}_{D^{\dagger}}(\eta)\hat{c}\hat{g}_{D^{\dagger}}(\eta) = \begin{bmatrix} c_{1,1} & c_{1,2} & \cdots & c_{1,D^{\dagger}-1} & \eta c_{1,D^{\dagger}} & \cdots & \eta c_{1,D} \\ c_{2,1} & c_{2,2} & \cdots & c_{1,D^{\dagger}-1} & \eta c_{1,D^{\dagger}} & \cdots & \eta c_{1,D} \\ \vdots & \vdots & \ddots & & \vdots & & \vdots \\ \vdots & \vdots & & \ddots & \vdots & & \vdots \\ \eta c_{D^{\dagger},1} & \eta c_{D^{\dagger},2} & \cdots & \eta c_{D^{\dagger},D^{\dagger}-1} & \eta^2 c_{D^{\dagger},D^{\dagger}} & \cdots & \eta^2 c_{D^{\dagger},D} \\ \vdots & \vdots & & \vdots & \vdots & \ddots & \vdots \\ \eta c_{D,1} & \eta c_{D,2} & \cdots & \eta c_{D,D^{\dagger}-1} & \eta^2 c_{D,D^{\dagger}} & \cdots & \eta^2 c_{D,D} \end{bmatrix} \tag{4}$$

so the elderly are more isolated among themselves, since $\eta < 1 \implies \eta^2 < \eta$, whilst still having reduced contacts with the other age classes.

Summarising the interventions implemented, we have:

- *Self-Isolation*: Self-isolation of not hospitalised symptomatic individuals, either being tested or not. It receives starting and ending date of intervention that models $\theta_{selfis}(t)$. Also receives coverage $selfis_{cov}$ and efficacy $selfis_{eff}$ values, giving $P_{selfis} = selfis_{cov} selfis_{eff} \theta_{selfis}(t)$;
- *Physical Distancing*: Reduction in contacts in other than home, school and work environments. It comprises interventions such as churches closure, maximum occupation in supermarkets and others. It receives starting and ending date of intervention, that models $\theta_{dist}(t)$. Also receives coverage $(dist_{cov})$ and efficacy $(dist_{eff})$ values;
- *Handwashing*: This intervention comprises individual protection measures, such as personal hygiene and mask usage. It receives starting and ending date of intervention, that models $\theta_{hand}(t)$. Also receives coverage $(hand_{cov})$ and efficacy $(hand_{eff})$;

- *Work from home*: This intervention models workers working from home. It receives starting and ending date of intervention, that models $\theta_{work}(t)$. Also receives coverage $(work_{cov})$ and efficacy $(work_{eff})$;
- *School closure*: This intervention models closing schools in the location. It receives starting and ending date of intervention, that models $\theta_{school}(t)$. Also receives coverage $(school_{cov})$ and efficacy $(school_{eff})$;
- *Cocoon elderly*: This intervention models the isolation of a proportion of the elderly population, given a minimum age $D^{\dagger}$. It receives starting and ending date of intervention, that models $\theta_{cocoon}(t)$. Also receives coverage $(cocoon_{cov})$ and efficacy $(cocoon_{eff})$.
- *Travel ban*: This intervention models the closure of frontiers of the location. It also models isolation at entrance of cases from outside. It receives the mean value of imported cases $(mean\_imports)$ and an efficacy value $(travel_{eff})$ of this restriction. This gives $imports = (1 - travel_{eff}) mean\_imports$, the value of new cases inserted by day in the model.

Taking into account that Self-Isolated $(\mathbf{X})$ individuals are only able to infect through home and "other" matrices, we can wrap everything in a force of infection given by:

$$\begin{aligned}\lambda &= (1 - hand(t)) p\bar{c}[\rho\mathbf{E} + \mathbf{I} + \mathbf{CL} + imports + \rho_s(\mathbf{H} + \mathbf{ICU} + \mathbf{ICUH})]/P \\ &\quad + (1 - hand(t)) p(\bar{c}_{home} + \bar{c}_{other})(\mathbf{X} + \mathbf{HC} + \mathbf{ICUC})/P \end{aligned} \tag{5}$$

where $hand(t) = hand_{cov} hand_{eff} \theta_{hand}(t)$.

#### C. Hospital burden

We slightly change the way hospital burden is added to the model from Aguas et al. (1). We assume that, if the occupation of beds is under some threshold value the health system infrastructure is able to handle correctly any new entrance to the hospital. After this threshold $q$, some of the patients might not find the needed support due to hospital overload, until full capacity, where patients are not accepted anymore. This is modelled by the following function:

$$f(x) = \begin{cases} 0, & \text{if } x < q, \\ 1 - (x(b - ax) + c), & \text{if } q \leq x \leq 1 \\ 1, & \text{if } x > 1 \end{cases} \tag{6}$$

Where $x$ is the ratio between number of patients and available beds and $a$, $b$, $c$ are computed in a way to ensure continuity of the function and its first derivative:

$$\begin{aligned} a &= \frac{1}{q(q-1) + (q^2 - 1)} \\ b &= 2aq \\ c &= a - b \end{aligned}$$

We have then $ICU_c$ being computed using $\mathbf{ICU}$/number of ICU beds, $H_c$ computed using $(\mathbf{ICUH} + \mathbf{H})$/number [illegible] (as $\mathbf{ICUH}$ uses common beds). Finally, to model priority of common beds to ICU needing individuals compared to $\mathbf{H}$ we simply assume that $ICU_h = H_c^2$, since these values are between 0 and 1, therefore $ICU_h \leq H_c$. The figure 1 shows these values assuming that $q = 0.65$.

### III. List of interventions

Here we show the list of interventions used as input of the model. Table III comprises all interventions used in the fitting of the standard model, which are equal to those used in the model with percolation. Figure 2 shows the timeline of these interventions. We also supplement with an additional table with the interventions used in the model with stronger interventions (Table IV).

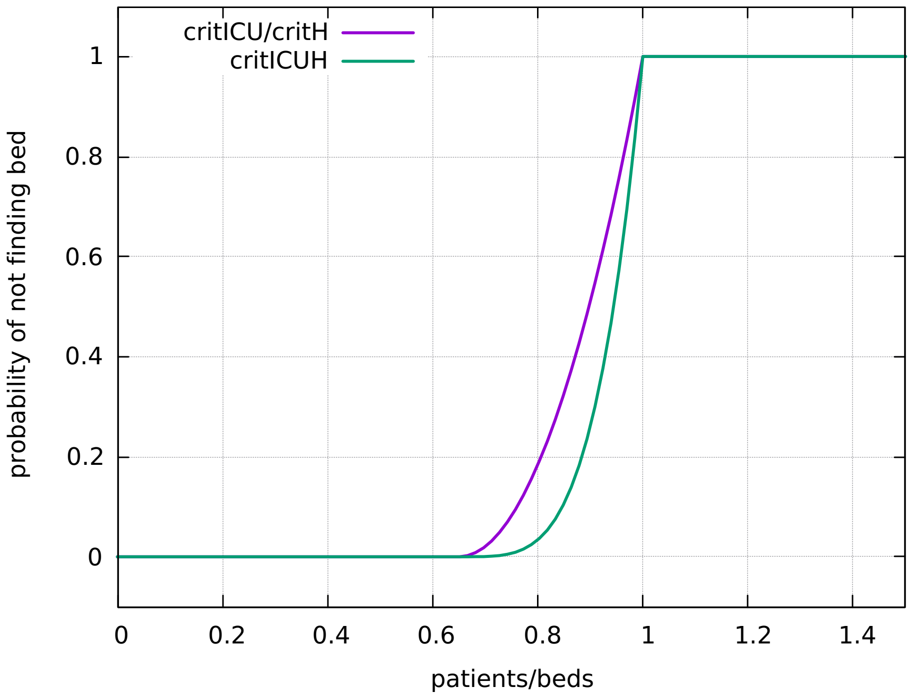


**Fig. 1:** Probability of not finding a bed as function of the current occupancy. $q = 0.65$.

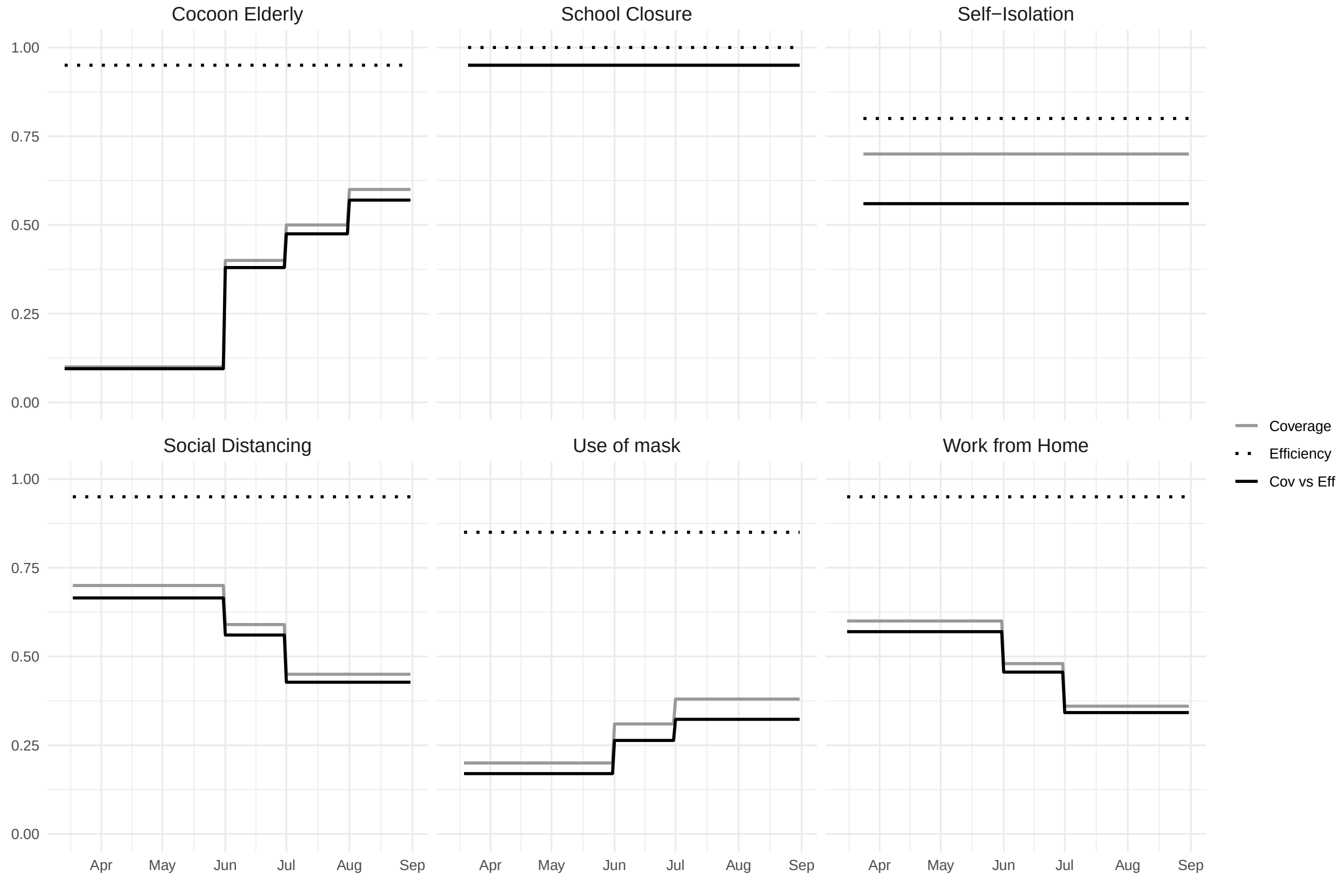


**Fig. 2:** Diagram of coverage, efficiency and their product for each of the considered non-pharmaceutical interventions considered in the standard model and the model with percolation. The diagrams for the standard model with 30% more NPI coverage would be similar, only30% higher values.

| Self Isolation | | | |
|---|---|---|---|
| Start date | End date | Coverage | Efficiency |
| 2020-03-24 | 2020-08-31 | 0.70 | 0.80 |
| **Social Distancing** | | | |
| Start date | End date | Coverage | Efficiency |
| 2020-03-18 | 2020-05-31 | 0.70 | 0.95 |
| 2020-06-01 | 2020-06-30 | 0.59 | 0.95 |
| 2020-07-01 | 2020-08-31 | 0.45 | 0.95 |
| **School Closure** | | | |
| Start date | End date | Coverage | Efficiency |
| 2020-03-21 | 2020-08-31 | 0.95 | 1.00 |
| **Use of Mask** | | | |
| Start date | End date | Coverage | Efficiency |
| 2020-03-19 | 2020-05-31 | 0.20 | 0.85 |
| 2020-06-01 | 2020-06-30 | 0.35 | 0.85 |
| 2020-07-01 | 2020-08-31 | 0.42 | 0.85 |
| **Work from Home** | | | |
| Start date | End date | Coverage | Efficiency |
| 2020-03-16 | 2020-05-31 | 0.60 | 0.95 |
| 2020-06-01 | 2020-06-30 | 0.48 | 0.95 |
| 2020-07-01 | 2020-08-31 | 0.36 | 0.95 |
| **Cocoon Elderly** | | | |
| Start date | End date | Coverage | Efficiency |
| 2020-03-14 | 2020-05-31 | 0.10 | 0.95 |
| 2020-06-01 | 2020-06-30 | 0.40 | 0.95 |
| 2020-07-01 | 2020-07-31 | 0.50 | 0.95 |
| 2020-08-01 | 2020-08-31 | 0.60 | 0.95 |
| **Travel Ban** | | | |
| Start date | End date | Mean imports | Efficiency |
| 2020-02-19 | 2020-03-18 | 0.20 | 0.0 |
| 2020-03-19 | 2020-08-31 | 0.20 | 0.70 |

**TABLE III:** List of interventions used for model fitting in the case of Standard and Percolation model.

### IV. Model Fitting

To fit the model onto epidemiological data, we used consolidated 2020 time series from Severe Acute Respiratory Infection (SARI) hospitalisations and deaths in São Paulo from the SIVEP-Gripe database (4) between 15 March and 31 August 2020. In Brazil, SARI case notification is compulsory (leading to high reporting rates) and SARS-CoV-2 is included as a SARI category. Though, due to the lack of extensive testing, we assume using only SARS-CoV-2 confirmed cases would lead to an underestimation of the actual number of cases. Hence, we assume that SARI cases are a better approximation to the number of SARS-CoV-2, rather than only cases confirmed by PCR tests. Also, note that this are severe cases and, therefore, hospitalised. Hence, we are fitting SARI cases to the sum over all hospitalised compartments of the model.

We specifically chose to use weekly time series for new cases and new deaths to avoid carrying past information into current values, as could happen if we used time series of cumulative data.

To perform a nonlinear least squares fitting of the free parameters ($p$, $T_{perc}$, $h_{steep}$, $startdate$) to the data, we used the Levenberg-Marquardt algorithm implemented in the minpack.lm R package (5).

| Self Isolation | | | |
|---|---|---|---|
| Start date | End date | Coverage | Efficiency |
| 2020-03-24 | 2020-08-31 | 0.91 | 0.80 |
| **Social Distancing** | | | |
| Start date | End date | Coverage | Efficiency |
| 2020-03-18 | 2020-05-31 | 0.91 | 0.95 |
| 2020-06-01 | 2020-06-30 | 0.767 | 0.95 |
| 2020-07-01 | 2020-08-31 | 0.585 | 0.95 |
| **School Closure** | | | |
| Start date | End date | Coverage | Efficiency |
| 2020-03-21 | 2020-08-31 | 1.00 | 1.00 |
| **Use of Mask** | | | |
| Start date | End date | Coverage | Efficiency |
| 2020-03-19 | 2020-05-31 | 0.20 | 0.85 |
| 2020-06-01 | 2020-06-30 | 0.35 | 0.85 |
| 2020-07-01 | 2020-08-31 | 0.42 | 0.85 |
| **Work from Home** | | | |
| Start date | End date | Coverage | Efficiency |
| 2020-03-16 | 2020-05-31 | 0.78 | 0.95 |
| 2020-06-01 | 2020-06-30 | 0.624 | 0.95 |
| 2020-07-01 | 2020-08-31 | 0.468 | 0.95 |
| **Cocoon Elderly** | | | |
| Start date | End date | Coverage | Efficiency |
| 2020-03-14 | 2020-05-31 | 0.13 | 0.95 |
| 2020-06-01 | 2020-06-30 | 0.52 | 0.95 |
| 2020-07-01 | 2020-07-31 | 0.65 | 0.95 |
| 2020-08-01 | 2020-08-31 | 0.78 | 0.95 |
| **Travel Ban** | | | |
| Start date | End date | Mean imports | Efficiency |
| 2020-02-19 | 2020-03-18 | 0.26 | 0.0 |
| 2020-03-19 | 2020-08-31 | 0.26 | 0.70 |

**TABLE IV:** List of interventions used for model fitting with stronger interventions.

In order to fit both new cases ($C$) and new deaths ($D$), we had to account for residuals in different scales. One way to do that was by normalising each of the variables in respect to their total sum. The resulting residual ($R$) is, therefore:

$$R = \frac{\sum(C_{model} - C_{observed})}{\sum C_{observed}} + \frac{\sum(D_{model} - D_{observed})}{\sum D_{observed}}$$

The algorithm minimise the square of this quantity, while evaluating the respective negative log-likelihood and minimising it.

To perform this kind of non-linear optimisation, we need to input the algorithm with a series of initial guesses. We tested a wide range of $startdate$ values (from $2020-10-01$ to $2020-02-24$) and for each one we ran the fitting algorithm using several reasonable initial guesses for the other free parameters. Hence, this method gives us fitted $p$, $T_{perc}$ and $h_{steep}$ for each $startdate$ considered.

With the goal to find a probability distribution for the fitted parameters (2), we selected the run which returned the lowest residual for each $startdate$, with its respective ($p$, $T_{perc}$, $h_{steep}$) set. We then computed the negative log-likelihood for each start date, $L_t$:

$$L_t = N \ln \left( \frac{1}{N} \sum_{i=1}^{N} R^2_{i,t}, \right)$$

from which we can therefore derive the probability for each startdate, given by

$$P_t = \frac{exp(-L_t + min(\{L_t\}))}{\sum_t exp(-L_t + min(\{L_t\}))}.$$

Finally, maximising the probability (equivalent to minimising the negative log-likelihood), we find sets of best fitted parameters for each of the model versions considered (See Table V).

### V. Model Comparison

In order to be able to compare the three different nested model versions plausibilities, we calculated their Akaike Information Criteria ($AIC$) (6). Using the results from the best fitting to data, for each model version $i$, the $AIC_i$ is

$$AIC_i = 2NLL_{min} + 2n_{par} \tag{7}$$

where $NLL_{min}$ is the negative log-likelihood from the best fitting run (which will be the minimum for that version) and $n_{par}$ is the number of fitted parameters for that version (which will be 2 for the standard models and 4 for the model with percolation).

To make the comparison even easier, we also calculate $AIC$ differences, defined as

$$\Delta AIC_i = AIC_i - AIC_{min} \tag{8}$$

where $AIC_{min}$ is the minimum over all AIC values obtained.

The resultant $AIC$ and $\Delta AIC$ obtained for each model version can be found in Table 1, in the main paper.

| Model | Parameter | Mean | SD | Quartile 2.5 | Quartile 50 | Quartile 97.5 |
|---|---|---|---|---|---|---|
| Percolation | startdate | 2020-01-30 | - | 2020-01-30 | 2020-01-30 | 2020-01-30 |
| Percolation | $p$ | 0.0461 | 0.0002 | 0.0461 | 0.0461 | 0.0462 |
| Percolation | $T_{perc}$ | 0.516 | 0.003 | 0.513 | 0.516 | 0.520 |
| Percolation | $h_{steep}$ | 4.83 | 0.02 | 4.81 | 4.83 | 4.84 |
| Standard | startdate | 2020-01-15 | - | 2020-01-15 | 2020-01-15 | 2020-01-15 |
| Standard | $p$ | 0.0294 | $\approx 0$ | 0.0294 | 0.0294 | 0.0294 |
| Standard + 30% NPIs | startdate | 2020-01-30 | - | 2020-01-30 | 2020-01-30 | 2020-02-01 |
| Standard + 30% NPIs | $p$ | 0.0402 | 0.0003 | 0.0401 | 0.0401 | 0.0411 |

**TABLE V:** List of parameters values for the three models with mean, standard deviation (SD), and 2.5$^{\text{th}}$, 50$^{\text{th}}$ and 97.5$^{\text{th}}$ percentiles.

### References

[1] R. Aguas, L. White, N. Hupert, R. Shretta, W. Pan-Ngum, O. Celhay, A. Moldokmatova, F. Arifi, A. Mirzazadeh, H. Sharifi, K. Adib, M. N. Sahak, C. Franco, and R. Coutinho. Modelling the COVID-19 pandemic in context: an international participatory approach. *BMJ Global Health*, 5(12), 2020. doi:10.1136/bmjgh-2020-003126. URL https://gh.bmj.com/content/5/12/e003126.

[2] K. P. Burnham and D. R. Anderson. *Model Selection and multi-model inference: A practical information-theoretic approach*. Springer New York, 2013.

[3] M. Cevik, M. Tate, O. Lloyd, A. E. Maraolo, J. Schafers, and A. Ho. SARS-CoV-2, SARS-CoV-1 and MERS-CoV viral load dynamics, duration of viral shedding and infectiousness – a living

systematic review and meta-analysis. July 2020. doi:10.1101/2020.07.25.20162107. URL https://doi.org/10.1101/2020.07.25.20162107.

[4] Datasus. Srag 2020 - banco de dados de síndrome respiratória aguda grave - incluindo dados da covid-19, 2020. URL https://opendatasus.saude.gov.br/dataset/bd-srag-2020.

[5] T. V. Elzhov, K. M. Mullen, A.-N. Spiess, and B. Bolker. *minpack.lm: R Interface to the Levenberg-Marquardt Nonlinear Least-Squares Algorithm Found in MINPACK, Plus Support for Bounds*, 2016. URL https://CRAN.R-project.org/package=minpack.lm. R package version 1.2-1.

[6] F. J. Fabozzi, S. M. Focardi, S. T. Rachev, and B. G. Arshanapalli. *Appendix E: Model Selection Criterion: AIC and BIC*, pages 399–403. John Wiley & Sons, Ltd, 2014. ISBN 9781118856406. doi:https://doi.org/10.1002/9781118856406.app5. URL https://onlinelibrary.wiley.com/doi/abs/10.1002/9781118856406.app5.

[7] IBGE. Tábuas completas de mortalidade, 2019. URL https://www.ibge.gov.br/estatisticas/sociais/populacao/9126-tabuas-completas-de-mortalidade.html?=&t=resultados.

[8] IBGE. Pesquisas estatísticas do registro civil - tabela 2679 - nascidos vivos, por ano de nascimento, idade da mãe na ocasião do parto, sexo e lugar do registro, 2021. URL https://sidra.ibge.gov.br/tabela/2679.

[9] T. P. Portella, S. R. Mortara, R. Lopes, A. Sánchez-Tapia, M. R. Donalísio, M. C. Castro, V. R. Venturieri, C. G. Estevam, A. F. Ribeiro, R. M. Coutinho, M. A. de Sousa Mascena Veras, P. I. Prado, and R. A. Kraenkel. Temporal and geographical variation of COVID-19 in-hospital fatality rate in brazil. Feb. 2021. doi:10.1101/2021.02.19.21251949. URL https://doi.org/10.1101/2021.02.19.21251949.

[10] H. Salje, C. T. Kiem, N. Lefrancq, N. Courtejoie, P. Bosetti, J. Paireau, A. Andronico, N. Hozé, J. Richet, C.-L. Dubost, Y. L. Strat, J. Lessler, D. Levy-Bruhl, A. Fontanet, L. Opatowski, P.-Y. Boelle, and S. Cauchemez. Estimating the burden of SARS-CoV-2 in france. *Science*, 369(6500): 208–211, May 2020. doi:10.1126/science.abc3517. URL https://doi.org/10.1126/science.abc3517.

[11] W. W. Sun, F. Ling, J. R. Pan, J. Cai, Z. P. Miao, S. L. Liu, W. Cheng, and E. F. Chen. Epidemiological characteristics of COVID-19 family clustering in Zhejiang Province. *Chinese journal of preventive medicine*, 54(6):625–629, 2020. ISSN 02539624. doi:10.3760/cma.j.cn112150-20200227-00199.

[12] W. E. Wei, Z. Li, C. J. Chiew, S. E. Yong, M. P. Toh, and V. J. Lee. Presymptomatic transmission of SARS-CoV-2 — singapore, january 23–march 16, 2020. *MMWR. Morbidity and Mortality Weekly Report*, 69(14):411–415, Apr. 2020. doi:10.15585/mmwr.mm6914e1. URL https://doi.org/10.15585/mmwr.mm6914e1.

[13] Y. Wei, L. Wei, Y. Liu, L. Huang, S. Shen, R. Zhang, J. Chen, Y. Zhao, H. Shen, and F. Chen. A systematic review and meta-analysis reveals long and dispersive incubation period of COVID-19. June 2020. doi:10.1101/2020.06.20.20134387. URL https://doi.org/10.1101/2020.06.20.20134387.
